# Weekly SARS-CoV-2 sentinel in primary schools, kindergartens and nurseries, June to November 2020, Germany

**DOI:** 10.1101/2021.01.22.21249971

**Authors:** Martin Hoch, Sebastian Vogel, Laura Kolberg, Elisabeth Dick, Volker Fingerle, Ute Eberle, Nikolaus Ackermann, Andreas Sing, Johannes Huebner, Anita Rack-Hoch, Tilmann Schober, Ulrich von Both

## Abstract

A 12-week sentinel programme monitored SARS-CoV-2 in primary schools, kindergartens and nurseries. Out of 3169 oropharyngeal swabs, only two tested positive on rRT-PCR while general incidence rates were surging. Thus, children attending respective institutions are not significantly contributing to the pandemic spread when appropriate infection control measures are in place.

## Background

Children have been disproportionately affected by public health measures in the current pandemic *(1, 2)*. In contrast to other age groups, children have shown lower rates of severe acute respiratory syndrome Coronavirus 2 (SARS-CoV-2) positive cases, lower risk for symptomatic acute coronavirus disease (COVID-19), a generally milder course of disease with the exception of some rare manifestations, and lower secondary attack rates *(3-5)*. Compared to teenagers, susceptibility to infection in one- to ten-year-olds is estimated to be lower. Accumulating evidence shows that, given limited infection control measures, SARS-CoV-2 may spread sustainably in secondary/high schools, but to a lesser degree in primary schools and nurseries *(3, 6, 7)*.

Closure of childcare facilities and schools has been shown to negatively affect children’s, teenagers’ and parents’ physical and emotional well-being. Hence, various expert groups called to avoid closure of these institutions *(8-10)*. Against the background of pre-symptomatic transmission found in adults it is critical to public health authorities to be able to rely on real-life data monitoring the number of asymptomatic yet infected children attending educational institutions (11, 12). Some studies have reported low number of infected cases in primary schools and childcare facilities but were conducted during a lockdown or semi lockdown period *(4, 13)*.

### The study

The “Münchner Virenwächter” study aimed for implementation of a real-time sentinel program in a representative number of five primary schools and five (six in phase 2) nurseries/kindergartens in Munich, Germany. It was intended to accomplish both a timely detection of infected cases and to offer an additional level of safety to participating institutions while in regular operating mode. The study span over two phases (Figure 1): Phase 1 (June 15^th^ until July 26^th^, 2020) and phase 2 (September 7^th^ until November 1^st^, 2020). Participating institutions were randomly selected and written informed consents obtained in the first week of each phase. In order to correct for underrepresentation of younger children (1-5 years), an additional nursery/kindergarten was included into phase 2. Testing for SARS-CoV-2 via real-time reverse transcription PCR (rRT-PCR) from oropharyngeal swabs was implemented with weekly samples obtained from randomly selected children (n = 20) and staff (n = 5) in each institution. Swabs were taken on-site by trained medical personnel and results were timely reported to each institution. For rRT-PCR, specimens were processed using the ampliCube Coronavirus SARS-CoV-2 (Mikrogen, Germany) on a Bio-Rad CFX96 Touch rRT-PCR Detection System (Bio-Rad, Germany). Single gene results were retested with Xpert Xpress SARS-CoV-2 (Cepheid, USA). SARS-CoV-2 IgG antibody screening was performed at three sequential time points on consented staff members of each institution using the Liaison SARS-CoV-2 S1/S2 IgG (Diasorin, Italy) system (Figure 1). Reactive results were confirmed via recomLine SARS-CoV-2 IgG Lineblot (Mikrogen, Germany). Antibody screening was complemented by obtaining a throat swab at the same time to exclude active infection. Institutions were asked to fill out a questionnaire assessing implementation of infection control measures for phase 1 and 2, respectively.

**Figure 1.**
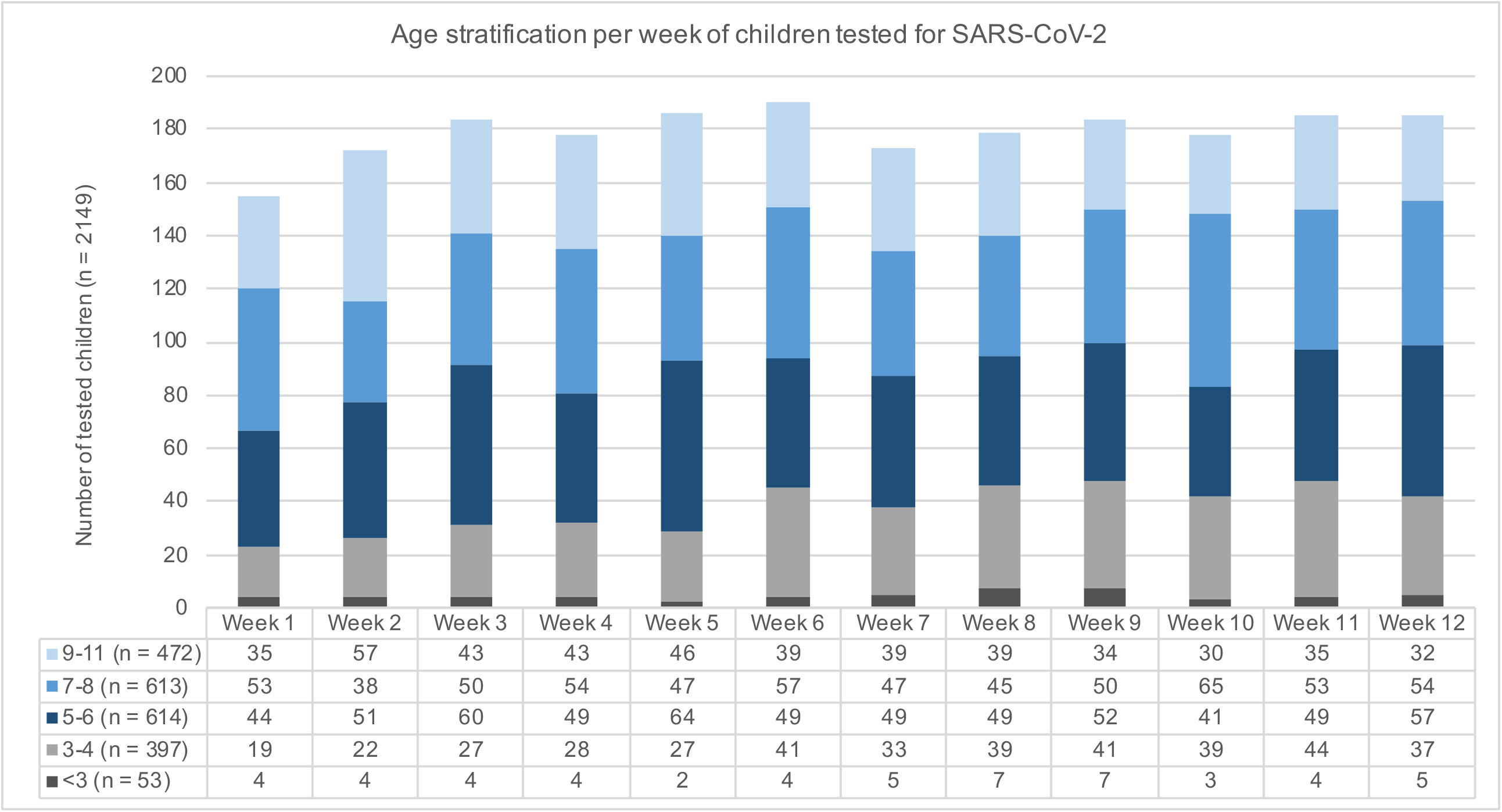
Timeline of Münchner Virenwächter study in context of pandemic activity in Munich, Germany. *7-day incidence rates derived from the national surveillance system according to the German infection protection act, Bavarian Health and Food Safety Authority as of November 28*^*th*^, *2020*.

In total 3169 individual oropharyngeal swabs were processed over the 12-week testing period with 2149 obtained from children (median age 7 years; range 1 - 11 years, male/female ratio 1.03) and 1020 from staff (median age 41 years; range 17-76 years, male/female ratio 0.13). In staff, 493 swabs were obtained during weekly testing and 527 to complement serology. A total of 527 blood samples from staff were subjected to SARS-CoV-2 IgG testing. Figure 2 illustrates pediatric sample distribution per study week. No SARS-CoV-2 infections were detected during phase 1 of the study. In phase 2 only week 12 yielded two test positive samples from one of the participating primary schools. All SARS-CoV-2 IgG samples from staff were test negative for timepoint 1 and 2; only one positive serologic result was detected at timepoint 3. In addition, we identified some changes for implemented infection control measures between study phase 1 and 2 and for individual facets between schools and childcare facilities, respectively (table 1). All children attending primary schools were wearing face masks on school premises, except when seated for teaching classes.

**Table 1.** Table. Comparing implementation of infection control measures in participating childcare facilities and primary schools.

| Infection control measures | Childcare facilities |  | Primary school |  |
| --- | --- | --- | --- | --- |
|  | Phase 1 | Phase2 | Phase 1 | Phase 2 |
| Reduced number of supervised children | 0/5 | 0/6 | 5/5 | 0/5 |
| Supervision of children by rotating groups/classes | 0/5 | 0/6 | 5/5 | 0/5 |
| Physical distancing between staff members inside | 5/5 | 6/6 | 5/5 | 5/5 |
| Physical distancing between staff members outside | 5/5 | 6/6 | 5/5 | 4/5 |
| Physical distancing between children inside | 1/5 | 2/6 | 5/5 | 5/5 |
| Physical distancing between children outside | 2/5 | 2/6 | 5/5 | 1/5 |
| Face mask for staff members inside | 0/5 | 6/6 | 4/5 | 5/5 |
| Face mask for staff members outside | 0/5 | 2/6 | 4/5 | 4/5 |
| Face mask for staff members during drop-off/collection of children | 3/5 | 6/6 | 4/5 | 5/5 |
| Face mask for parents during drop-off/collection of children | 5/5 | 6/6 | 4/5 | 5/5 |
| Parents allowed to enter premises when dropping or collecting kids | 5/5 | 4/6 | 1/5 | 2/5 |
| Washing hands before collection of children by parents | 3/5 | 5/6 | 1/5 | 1/5 |
| Use of bathroom facilities separate for individual groups/classes | 5/5 | 5/6 | 2/5 | 2/5 |
| Closure of garden/playground areas | 0/5 | 0/6 | 0/5 | 0/5 |
| Use of garden/playground areas separate for individual groups/classes | 4/5 | 4/6 | 5/5 | 5/5 |
| Handwashing before meals | 5/5 | 6/6 | 5/5 | 5/5 |
| Handwashing before entering classes / groups | 5/5 | 5/5 | 5/5 | 5/5 |
| Hand disinfectant dispensers provided on premises | 4/5 | 6/6 | 4/5 | 3/5 |
| Cancellation of common activities | 5/5 | 6/6 | 5/5 | 5/5 |

**Figure 2.**
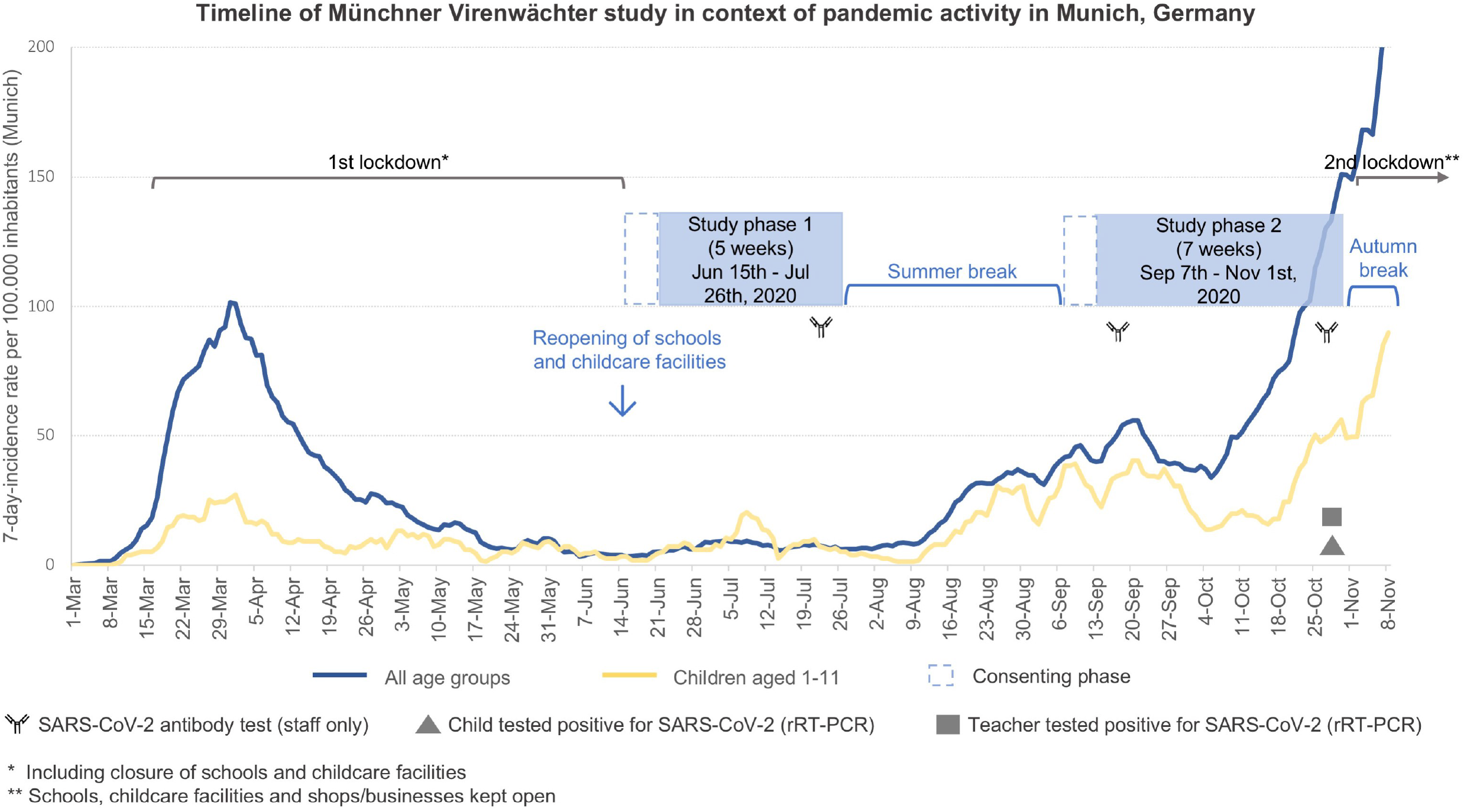
Distribution of weekly pediatric oropharyngeal swab samples for SARS-CoV-2 rRT-PCR testing; color code indicating individual age groups.

Designed during the first lockdown in Munich, our study intended to implement a feasible SARS-CoV-2 sentinel programme in primary schools and childcare facilities in anticipation of a second pandemic wave and rising incidence rates. While public health and political authorities were worried that childcare institutions would be significant drivers of the pandemic, our results suggest that this was not the case. While the study was not powered to accurately illustrate changes in incidences during low-incidence periods, we were able to detect 2 cases in a primary school, one child and his/her teacher during a high local seven-day incidence rate of 50/100.000 (children aged 1-11) and 150/100.000 (general population). During testing of 36 close contacts (33 classmates, 3 private contacts) only 1 additional case was identified in another asymptomatic child of the same class. Upon phone-interview-based contact tracing the teacher reported to have experienced unspecific symptoms of headache and malaise 6 days prior to testing. Thus, it seems reasonable to deduct that transmission occurred from staff to both children.

## Conclusions

A few reports have assessed the role of children in the dynamics of SARS-CoV-2 transmission. A recent study conducted in day-care centers using buccal mucosal and anal swabs for SARS-CoV-2 detection concluded that shedding of the virus was very rare and that day-care centers are no relevant reservoir in a low prevalence setting *(14)*. However, this study used self-testing, lacked pharyngeal swabs and was conducted during a minimal local incidence rate. Our study covered both a low and high 7-day incidence period using oropharyngeal swabs on children aged 1 to 11 years. Ehrhardt and colleagues demonstrated very low transmission in schools and childcare facilities. While this is in line with our findings, again this study was conducted during a period of low infection activity and partial lockdown; furthermore, individuals up to 19 years of age were included – representing a population with a very different known epidemiologic relevance in this pandemic *(4)*. Finally, Ismail and colleagues presented complementing data from the UK showing that staff members had an increased risk of SARS-CoV-2 infection compared to students in any educational setting and that the majority of cases linked to outbreaks were in staff. Secondary attack rate analysis of the cases found in our study also suggests that infection occurred from staff to children (15). In addition, low prevalence for SARS-CoV-2 antibodies in staff over the three-months study period suggests no relevant infection activity in neither work nor private setting.

Feedback received from participating institutions was unequivocally positive. Having a weekly, easy-to-integrate sentinel running rendered an important additional level of safety to daily routine. Consequently, timely communication of test results is likely to have strengthened adherence to infection control measures. It is likely that children reflect infection activity of the local community. But we conclude that asymptomatic children attending primary schools, kindergartens and nurseries are not significantly contributing to pandemic distribution of SARS-CoV-2 while adhering to infection control measures described above, even during high local background incidence. Thus, they are unlikely to initiate clusters or outbreaks in the community when these institutions continue to play their critical role for the physical and emotional well-being of children and their families.

## Data Availability

All data presented in this manuscript will be available from the authors upon request.

## Acknowledgements

Authors would like to thank Annalena Branz, Felix Flachenecker, Janina Ludwig, Adrian Rödig, Maria-Sophia Stadler, Jasmin Mahdawi and Johannes Nowak for helping in the field. Authors would like to acknowledge Rüdiger von Kries for statistical advice and thank all participating institutions, their staff as well as all children and their parents for their valuable support. Authors would also like to thank the *Bayerisches Staatsministerium für Unterricht und Kultus, the Referat für Bildung und Sport der Landeshauptstadt München and the Referat für Gesundheit und Umwelt (RGU)* for supporting our study. No external funding was received for this work. This study was approved by the ethics committee of the Ludwig-Maximilians University (LUM) under project number 20-484.

## Notes

### Competing Interest Statement

The authors have declared no competing interest.

### Funding Statement

This study did not receive external funding.

### Author Declarations

This study was approved by the ethics committee of the Ludwig-Maximilians University (LUM) under project number 20-484.

